# The test-retest reliability and agreement between a fixed frame and belt-stabilised handheld dynamometer for isometric hip flexion and extension peak force measurement in recreational cyclists

**DOI:** 10.1101/2025.06.29.25330525

**Authors:** Dion D’Mello, Benn Digweed, Tom Hughes

**Affiliations:** Department of Health Professions, Manchester Metropolitan University, Manchester, UK; United Kingdom Sports Institute, UK Sports Institute High Performance Centre, Manchester Institute of Health and Performance, Manchester, UK; Institute of Sport, Manchester Metropolitan University, Manchester, UK

## Abstract

**Introduction:** Cycling performance is influenced by hip flexor and extensor muscle strength. While belt stabilised handheld dynamometers (B-HHD) are valid for measuring isometric hip muscle strength, fixed frame dynamometers are becoming popular, offering potentially better stability and reliability. However, the reliability of both devices has not been examined in cyclists. This study evaluated the test-retest reliability and agreement between a B-HHD (MicroFET2, Hoggan Scientific) and a fixed-frame dynamometer (ForceFrame (FF) Max, Vald Performance) for hip flexion and extension peak force measurement in cyclists.

**Methods:** A test-retest design was used. Twenty-five recreational cyclists (age ± SD: 36.64 (±12.34) years; 22 males) were tested twice, approximately 72 hours apart. Three unilateral maximal voluntary isometric contractions (MVIC) of the hip flexors and extensors of each limb were performed, using the B-HHD and FF in a random order. Within and between session reliability was determined using Intraclass correlation coefficients _3.1 & 3.k._ Standard error of measurements (SEM) and minimal detectable changes (MDC) were calculated. Agreement was assessed using 95% limits of agreement (LOA).

**Results:** For hip flexion, within and between session reliability was good to excellent, and SEMs were similar (B-HHD ICCs = 0.77-0.93, SEMs = 14.25-22.71N (7.19-10.38%); FF ICCs = 0.77-0.95, SEMs = 7.80N-18.98N (3.47%-8.54%)). FF MDCs were lower within-session (21.61-39.48N (9.60-17.97%)) than B-HHD MDCs (39.50-62.95N (19.94-28.78%)), but similar between-sessions (FF MDCs= 41.25-52.61N (19.42-23.66%); B-HHD MDCs 41.21N-48.95N (18.53-23.77%)).

For hip extension, both devices demonstrated good to excellent reliability and SEMs were similar (B-HHD ICCs = 0.90-0.95, SEMs = 15.77-21.53N ( 7.38-9.96%); FF ICCs = 0.85-0.95, SEMs =19.21-29.05N (7.82-11.78%) within and between sessions). All LOA exceeded a 20N acceptability threshold.

**Conclusion:** Both devices are reliable in recreational cyclists, but large MDCs suggest that caution is needed when interpreting repeated measurements. Both devices cannot be used interchangeably due to poor agreement.

## Introduction

Cycling places physiological stress on the metabolic, cardiovascular and musculoskeletal systems, which can improve cardiovascular fitness (1), cognitive function and overall wellbeing (2). It is associated with a reduced risk of cardiovascular disease (CVD) and related risk factors (including obesity, hypertension and elevated trigylcerides) (3). Cycling is linked to a 17% reduction in CVD mortality (3) and a 20% reduction in all-cause mortality (1), making it a highly recommended activity for public health improvement (4). Additionally, cycling has become increasingly popular in the United States (US) (5, 6) and other European countries (7, 8) for transportation, recreational (5) and competitive purposes (6).

Approximately 85% of cycling power is generated during the pedal downstroke (9) by the hip extensor, knee extensor, and ankle plantarflexor muscles (10). During pedal upstroke, power is generated by the hip and knee flexors, with biarticular muscles maintaining joint stability (11). To enhance muscular strength and cycling performance, recreational and competitive cyclists commonly combine resistance training with cycling endurance training (12). Indeed, resistance training has been shown to improve peak power output in competitive cyclists (13, 14), cycling economy and VO_2_ Max in moderately trained cyclists (15, 16) and pedalling cadence efficiency in recreational athletes (14, 16, 17). However, these latter benefits are less clear in highly trained cyclists (12, 14–16).

Given the importance of hip strength in cycling performance and the risk of injury associated with cycling participation (18), regular muscle strength assessments may provide valuable data for cyclists, coaches and health professionals. Indeed, regular physical testing can be useful for performance tracking, training evaluation and planning (19–21), setting rehabilitation targets (19) and assessing treatment effectiveness following injury (22).

Isokinetic dynamometers (IKD) are considered the gold standard for muscle strength assessment due to their reliability and ability to measure various strength components (including peak force, work, power, endurance and force curves) (22). However, IKDs are expensive, non-portable and require expert training, limiting their use to clinical (23, 24) and research settings. In contrast, portable and simple hand-held dynamometers (HHDs) (such as the MicroFET2, Hoggan Scientific, Salt Lake City, USA) are commonly used to measure isometric strength, due to their cost-effectiveness, mass-testing capabilities (25), accessibility, and ease of use without specialist training (26). HHDs can quantify maximal isometric voluntary contractions (MVICs), which are valid assessments of maximal strength (21).

HHDs show acceptable intra and inter-rater reliability for measuring isometric hip flexion and extension MVICs (weighted correlation coefficient (WCC) ranges: 0.79-0.91 and 0.81-0.87, respectively) (27). However, measurements can be influenced by the examiner’s strength, potentially causing inter-rater bias (28), and the lack of device stabilisation can introduce errors (29). External fixation methods, such as belt stabilisation, can eliminate examiner-provided resistance and improve test-retest reliability (WCC ranges = 0.91-0.93, ICC _2.1_ = 0.91-0.95) (27, 30). Importantly, population characteristics influence device reliability (31, 32), but to date, the reliability of external, belt-stabilised HHDs (B-HHDs) has not been investigated in cyclists, so it is unclear if these estimates apply to this population.

While belt stabilisation may improve reliability, newer fixed frame dynamometers (such as the Vald ForceFrame (herein termed FF) Max (Vald Performance, Brisbane, Australia)), may offer superior stabilisation because sensors are mounted directly to a frame and base. Fixed frame devices are also highly adjustable, which allows testing of most muscle groups. A different iteration of the FF (i.e. ForceFrame Fold) has shown good to excellent test-retest reliability for hip adduction (ICC_2.1_= 0.81-0.90) and moderate to excellent for hip abduction MVICs (ICC _2.1_= 0.72-0.92) in field sport athletes (33). However, to the best of our knowledge, no previous studies have investigated the reliability of any FF version for measuring hip flexion and extension MVICs in any population.

Given the growing preference to replace HHDs with fixed frame devices, (34) it is essential to determine whether measurements from both devices can be interchanged. This may help to ensure consistent and accurate data in settings where both devices are available, where longitudinal data have already been collected by B-HHDs, or when practitioners are deciding whether to use one method over another.

The aim of this study was to evaluate the test-retest reliability and agreement between a belt-stabilised MicroFET2 HHD and the ForceFrame Max, for measurment of hip flexion and extension MVIC peak force in recreational cyclists.

## Materials and methods

This study was reported according to The Guidelines for Reporting Reliability and Agreement Studies (32) and the Consensus-based Standards for the Selection of health Measurement INstruments (COSMIN) Reporting Guideline for Studies on Measurement Properties of Patient Reported Outcomes Measures (35). The methodology has been detailed *a priori*, in a protocol available on Open Science Framework (36).

### Study design and setting

Since both devices were stabilised, examiner effects were neglible, so a test-retest design was utilised. Data were collected between 04/03/2024 and 22/03/2024 in a laboratory at Manchester Metropolitan University. The examiner was a Physiotherapy MSc student (36), with 3 years experience of using HHDs but no prior experience with the ForceFrame Max. Ten hours of pilot testing was completed for protocol refinement and familiarisation.

### Participants

Recreational and competitive cyclists were recruited using purposive sampling. Posters were placed in cycle shelters at Manchester Metropolitan University and distributed to three cycling clubs in the Greater Manchester area. Potential participants voluntarily contacted the lead researcher for eligibility confirmation and study details.

Participants were included if they were aged >18 years and completed > 150 minutes cycling per week at ‘vigorous’intensity ( > 10 miles per hour as defined by the Centre for Disease Control and Prevention (*37*)).

Participants were excluded if they: had an active neurological, cardiovascular or respiratory disease; were pregnant; had any lacerations, abrasions, or contusions at the distal femur affecting the area of HHD and FF load cell sensor application during testing; reported any musculoskeletal injury or orthopaedic surgery to the lumbar spine, pelvis or lower limbs or suffered any systemic illness within 3 months of the first assessment day.

Ethical approval was granted by the Faculty of Health and Education Research Ethics and Governance Committee at Manchester Metropolitan University (no.59674). All participants provided written informed consent prior to participation.

### Sample size

Sample size was calculated using the confidence interval (CI) width procedure of the ICC & SEM Power Sample Size Decision Assistant (https://iriseekhout.shinyapps.io/ICCpower) (*38*). Previously reported intraclass correlation coefficents (ICCs) for hip flexion and extension strength measurements were 0.91 and 0.90 for HHDs with external stabilisation (27) and 0.77-0.86 for the Groinbar, (Vald Performance, Brisbane, Australia) (23) a similar fixed frame device to the Forceframe Max used in this study. For a two-way mixed effects ICC model needed for a test-retest design, our calculation assumed a conservative expected correlation of 0.80, a moderate variance of 10 for between-participant test scores with no expected systematic differences, and aimed for a target CI width of 0.30. Our calculation indicated that recruitment of 25 participants would enable ICC estimation with CI precision of 0.26 for within-session analyses (three trials per testing session), and 0.33 for between-session analyses (mean of three trials are compared over two testing sessions) (see S1 Appendix for full calculation).

### Instrumentation and measurement parameters

For all participants, self-reported weekly cycling activity level (minutes) and the usual mode of cycling participation were collected. Baseline measurements of standing height (centimetres) and body mass (kilograms) were recorded on the first testing day using the Leicester Height Measure (Seca Ltd., Birmingham, UK) and the Personal Floor Scale MPE 250K100HM (Kern & Sohn, Germany).

A belt-stabilised Micro-FET2HHD (B-HHD), and ForceFrame Max Strength Testing System (FF) were used to assess hip flexion and hip extension peak force during MVICs, recorded directly into a laptop computer. B-HHD measurements were entered into an Excel (Microsoft, Washington, USA) spreadsheet by the examiner using a wired connection. The B-HHD was calibrated at the start of each testing day. The FF was zeroed between each test. Peak force from all MVICs were recorded in newtons (N) and retained on their continuous scale for analyses.

### Experimental procedure

All data were collected by the primary researcher who was unblinded. Before each testing session, participants completed a standardised 5-minute warm-up on a stationary bicycle at a self-determined moderate intensity. Across both testing days, participants completed unliateral hip strength assessments with the B-HHD and FF, for both limbs. To eliminate order effects, tests were randomised by muscle group, devices and limb, using the RAND function in Excel, conducted by an independent person.

For B-HHD hip flexion, participants were seated upright on an examination table with their arms crossed against their chest. For FF hip flexion, due to device constraints, testing was conducted with participants in supine with arms crossed against their chest. The examiner used a goniometer to ensure the hip was positioned to 90° flexion for both devices. The sensor for each device was positioned along the distal femur, 1 inch superior to the patellar base.

For B-HHD and FF extension, participants lay prone with their hands under their forehead. The examiner used a goniometer to ensure the knee tested limb was was flexed to 90° for both devices. The sensor of each device was positioned along the distal femur, 1 inch superior to the popliteal crease. All testing positions are presented in S2 Appendix.

For the FF tests, the crossbar height was reproduced on the second testing day. For all B-HHD tests, the fixation belt was secured to the dynamometer and examination table. The examiner manually stabilised the B-HHD to eliminate slipping but did not exert any downward pressure on the sensor.

For each muscle group, device and limb participants completed three practice trials at 50%, 70% and 90% of their self-perceived maximal efforts. Following these practice attempts, participants performed three 5-second MVICs. For each trial, the examiner provided a standardised instruction of “Begin now – 3, 2, 1 - GO - 1, 2, 3, 4, 5 - relax”. A 10-second rest period was given between trials and a 2-minute rest periods were provided upon test completion to minimise fatigue and allow for repositioning.

Trials were considered invalid and repeated if: participants altered their test position; the B-HHD slipped; an error was detected on the VALD interface. Participants were withdrawn if they experienced pain in their lower limbs or lumbar spine during testing. Upon completion, data collection was repeated at 72 hours later, or as close to 72 hours as possible if participants could not attend.

### Statistical analyses

Descriptive statistics (mean ± standard deviation (SD)) were calculated for height, body mass, cycling volume and age. Proportions were calculated for biological sex, and cycling modes.

All analyses were based on the assumption of normally distributed data. For within-session analyses, peak force (N) was compared across all three trials for each device, muscle group and limb on each testing day. For between-session analyses, mean peak force was calculated using all three trials for each participant according to device, muscle group, limb and testing day. Mean values were then compared across testing days.

Within session ICCs_3.1_ and between session ICCs_3.k_ were calculated with corresponding 95% CIs and interpreted as poor < 0.40; fair= 0.40-0.70; good=0.70-0.90; excellent > 0.90 (39). Standard error of measurements (SEM) were calculated using the formula 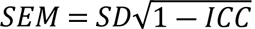. Minimal detectable changes (MDC) were calculated using the formula: *MDC* = *SEM* × 1.96 × √2 (40). SEM and MDC values were reported in newtons (N) and as percentages of mean values.

To determine the within-session agreement, the distribution of between device differences was assessed using quantile-quantile plots according to muscle group tested, limb and testing day. Bland-Altman plots with with 95% limits of agreement (LOA) were constructed (41). Previously, the MicroFET2 HHD has been shown to have concurrent validity with a similar fixed frame dynamometer system (KangaTech KT360, KangaTech, Melbourne, Australia), with a reported absolute mean difference of 7.74N (95%CI=1.86-16.40N) (42). Therefore, agreement was considered clinically acceptable if the 95%LOA were within 20N. All statistical analyses were initially conducted by DD using SPSS 29 (IBM, New York, USA), and repeated by TH using STATA 18 (StataCorp LLC, Texas, USA) for accuracy.

## Results

### Study Participation and missing data

Of twenty-six participants recruited, one withdrew due to pain during testing and was excluded from the analysis. The remaining participants included 22 males and 3 females with the following characteristics (mean ± SD): age 36.64 (±12.34) years; height 180.36 (±7.76) cm; mass 80.25 (± 9.86) kg; cycling activity 294 (± 183.24) minutes per week. Ten participants (40%) primarily cycled for travel, 10 (40%) were recreational road cyclists, 3 (12%) mountain bikers and 2 (8%) used a stationary cycle at the gym.

### Within-session Reliability

For the within-session analyses, descriptive statistics are presented in Table 1, and ICCs, 95% CIs, SEMs and MDCs are presented in Table 2.

**Table 1:** Within-session descriptive statistics.

| Within- session<br>Descriptive Statistics |  | Session 1 |  |  |  | Session 2 |  |  |  |
| --- | --- | --- | --- | --- | --- | --- | --- | --- | --- |
|  |  | Left limb |  | Right limb |  | Left Limb |  | Right limb |  |
| Measure | System | Mean | SD | Mean | SD | Mean | SD | Mean | SD |
| Hip<br>Flexion | B-HHD | 198.12 | 47.13 | 218.75 | 61.96 | 213.81 | 36.24 | 225.96 | 53.27 |
|  | FF | 206.33 | 39.09 | 219.67 | 45.92 | 218.47 | 35.10 | 225.08 | 35.17 |
| Hip<br>Extension | B-HHD | 206.19 | 69.35 | 214.86 | 66.79 | 213.67 | 70.67 | 217.28 | 73.03 |
|  | FF | 251.57 | 85.00 | 243.87 | 70.70 | 241.71 | 66.23 | 247.56 | 76.15 |

*Key: B-HHD = belt stabilised handheld dynamometer; FF = Forceframe; SD = standard*
*deviation*

**Table 2:**
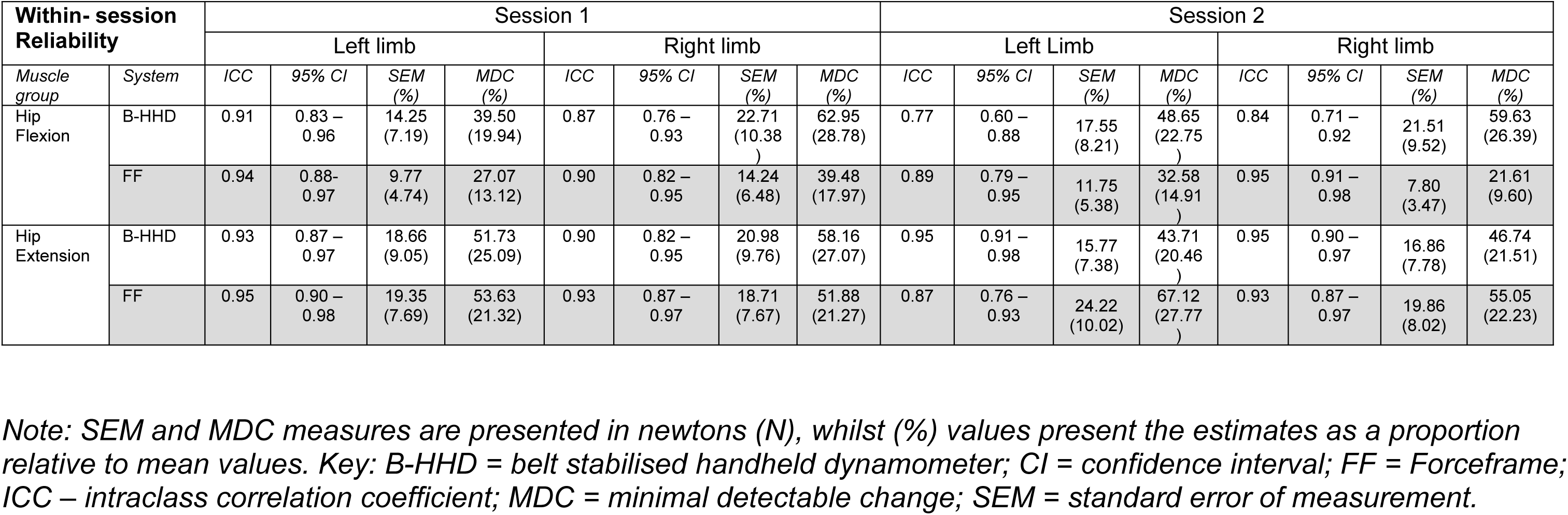
Within-session ICCs_3.1,_ 95%CIs, SEM and MDC estimates.

For hip flexion, both devices generally showed comparable good to excellent reliability and similar SEMs. Across both days and limbs, FF ICCs_3.1_ ranged between 0.89-0.95. B-HHD reliability of the the left limb on day 2 was slightly lower (ICC_3.1_=0.77), but all other values were similar to the FF (ICC_3.1=_0.84-0.91). FF SEMs were between 7.80 – 14.24N (or 3.47-6.48%). Similarly, B-HHD SEMs were between 14.25-22.71N (or 7.19-10.38%). However, FF MDCs were consistently lower (21.61-39.48N (9.60-17.97%)) than B-HHD MDCs (39.50-62.95N (19.94-28.78 %)).

For hip extension, both devices showed good to excellent reliability with similar SEM and MDC estimates. Across both days and limbs, FF ICCs_3.1_ were 0.87-0.95 and B-HHD ICCs_3.1_ were 0.90-0.95. FF SEMs were between 18.71-24.22N (7.67-10.02%), and B-HHD SEMs were 15.77 - 20.98N (7.38-9.76%). FF MDCs were between 51.88 – 67.12N (21.27-27.77%) and B-HHD MDCs were 43.71-58.16N (20.46-27.07%).

### Between-session reliability

Descriptive statistics, ICCs_3.k_, 95% CIs, SEMs and MDCs are presented in Table 3.

**Table 3:** Between-session descriptive statistics, ICCs, 95%CIs, SEM and MDC estimates.

| Between-session Reliability |  | Left limb |  |  |  |  |  | Right limb |  |  |  |  |  |
| --- | --- | --- | --- | --- | --- | --- | --- | --- | --- | --- | --- | --- | --- |
| Measure | System | Mean | SD | ICC | 95% CI | SEM (%) | MDC (%) | Mean | SD | ICC | 95% CI | SEM (%) | MDC (%) |
| Hip Flexion | B-HHD | 205.96 | 40.80 | 0.81 | 0.53 – 0.92 | 17.66 (8.57) | 48.95 (23.77) | 222.35 | 55.08 | 0.93 | 0.84 – 0.97 | 14.87 (6.69) | 41.21 (18.53) |
|  | FF | 212.40 | 36.68 | 0.84 | 0.61 – 0.93 | 14.88 (7.01) | 41.25 (19.42) | 222.37 | 40.00 | 0.77 | 0.49 – 0.90 | 18.98 (8.54) | 52.61 (23.66) |
| Hip Extension | B-HHD | 209.93 | 68.85 | 0.92 | 0.82 – 0.97 | 19.48 (9.28) | 54.01 (25.73) | 216.07 | 68.41 | 0.90 | 0.78 – 0.96 | 21.53 (9.96) | 59.67 (27.62) |
|  | FF | 246.64 | 74.39 | 0.85 | 0.66 – 0.93 | 29.05 (11.78) | 80.51 (32.64) | 245.71 | 71.97 | 0.93 | 0.84 – 0.97 | 19.21 (7.82) | 53.26 (21.68) |

*Note: SEM and MDC measures are presented in newtons (N), whilst (%) values present the estimates as a proportion relative to mean values. Key: B-HHD = belt stabilised handheld dynamometer; CI = confidence interval; FF = Forceframe; ICC – intraclass correlation coefficient; MDC = minimal detectable change; SEM = standard error of measurement.*

For hip flexion, FF reliability was consistently good (ICC_3.k._ range = 0.77-0.84). B-HHD reliability was good on the left limb (ICC_3.k=_ 0.81) and excellent on the right (ICC_3.k_ = 0.93). For both devices, SEMs and MDCs were similar (SEMs: FF = 14.88-18.98N (7.01-8.54%), B-HHD =14.87-17.66N (6.69-8.57%); MDCs: FF = 41.25-52.61N (19.42-23.66%), B-HHD 41.21N-48.95N (18.53-23.77%)).

For hip extension, both devices had good to excellent reliability across limbs (FF ICC_3.k_ = 0.85-0.93, B-HHD ICC_3.k_ = 0.90-0.92). SEMs and MDCs were also comparable (SEM: FF= 19.21 – 29.05N (7.82-11.78%), B-HHD = 19.48-21.53N (9.28-9.96%); MDCs: FF = 53.26 – 80.51N (21.68 – 32.64%), B-HHD 54.01-59.67N (25.73-27.62%)).

### Between-system agreement

The distribution of between-device differences was approximately normal across limbs, testing days and muscle groups (S3 Appendix). Bland and Altman plots are presented for hip flexion (Fig. 1) and extension (Fig.2). All estimates for mean differences and LOA are presented in S4 Appendix.

For hip flexion, mean agreement ranged from −0.88 to 8.22 N across limbs and testing days. For hip extension, the FF consistently produced higher peak force measures (mean agreement range = 28.04 to 45.38 N) across both limbs and days. All 95% LOAs were very wide, exceeding the 20N acceptable threshold, ranging from 126.98N (Fig. 1, plot C) to 289.09N (Fig.2, plot D).

**Fig.1.**
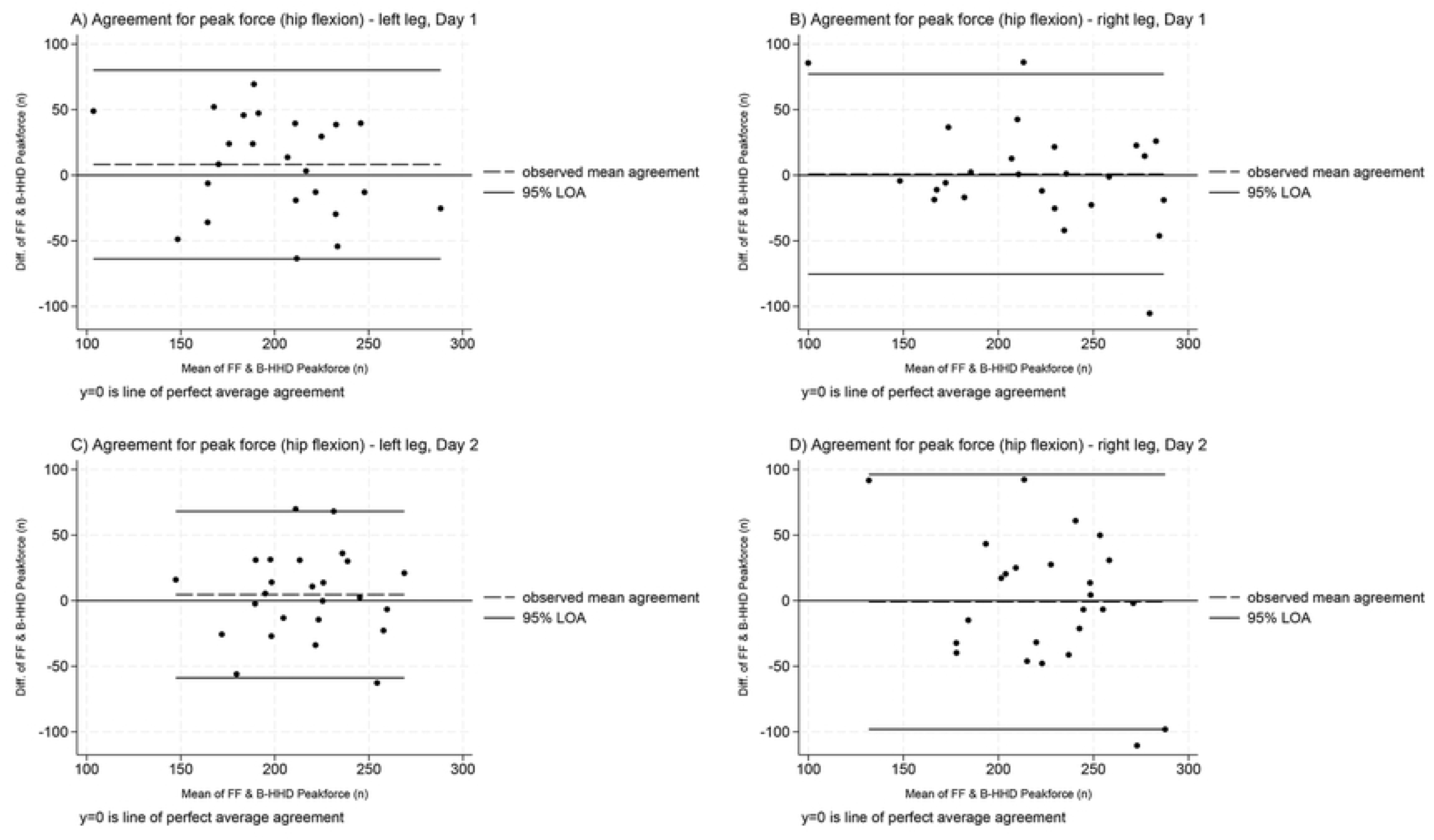
– Within-session agreement between devices for hip flexion peak force measurement (both limbs and testing days) *Note: The Y axis (difference between devices) at 0 indicates perfect agreement; positive values show the Forceframe recorded greater values, whereas negative values show the Belt-stabilised handheld dynamometer recorded greater values. Thick black lines correspond to 95% limits of agreement, dashed black lines represent observed mean agreement. Key: B-HHD = Belt-stabilised handheld dynamometer; diff = difference, FF= Forceframe; LOA = limits of agreement; n= newtons*.

**Fig.2.**
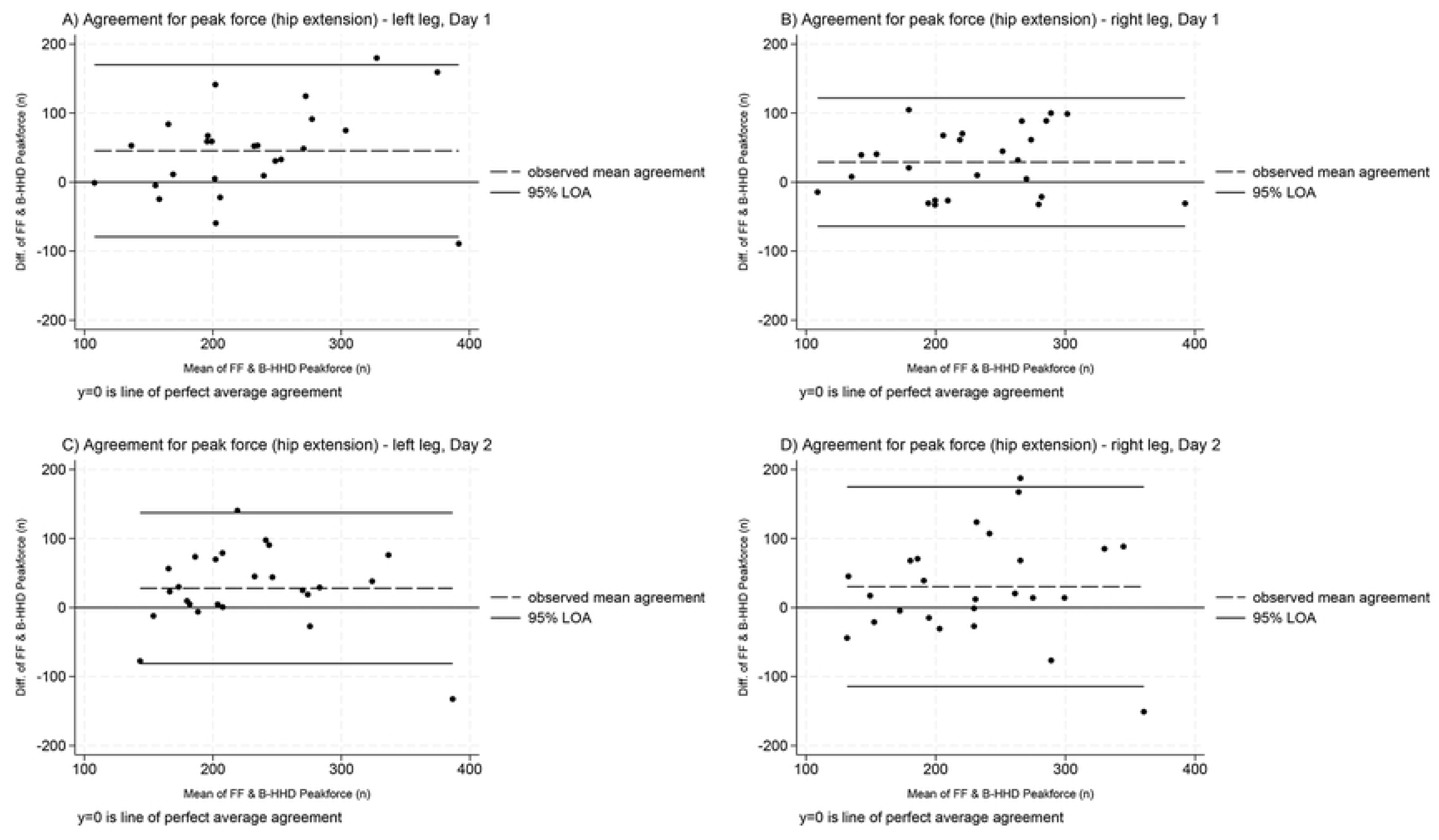
– Within-session agreement between devices for hip extension peak force measurement (both limbs and testing days) *Note: The Y axis (difference between devices) at 0 indicates perfect agreement; positive values show the Forceframe recorded greater values, whereas negative values show the Belt-stabilised handheld dynamometer recorded greater values. Thick black lines correspond to 95% limits of agreement, dashed black lines represent observed mean agreement. Key: B-HHD = Belt-stabilised handheld dynamometer; diff = difference, FF= Forceframe; LOA = limits of agreement; n= newtons*.

## Discussion

This study is the first to assess the test-retest reliability and agreement between a belt-stabilised, handheld MicroFET2 dynamometer and the Forceframe Max system for peak force hip flexion and extension measurements in cyclists, so offers novel data. While both devices initially appear reliable, the observed MDC and agreement estimates have important practical and clinical implications.

### Reliability and measurement error

For hip flexion, both devices demonstrated good to excellent within and between-session reliability and measurement error. Specifically, B-HHD ICCs ranged between 0.77-0.93, with SEMs of 14.25N-22.71N (7.19%-10.38%). These values are generally comparable to a recent meta-analysis of portable, externally stabilised dynamometers for seated hip flexion MVIC assessments, which reported intra-rater WCCs of 0.91 (95%CI=0.84-0.99) and average SEMs of 15.00N. This similarity was expected, as most devices in the meta-analysis used similar B-HHDs test positions and protocols to our study.

We also found that between-session B-HHD estimates (ICC _3.k_ =0.81-0.93) were comparable to those recently reported for peak torque by McNabb et al (ICC_2.1_ = 0.91) (30). However, our B-HHD SEMs (6.69%-8.57%) indicated greater precision than the 14% reported by McNabb et al (30). While both studies used similar protocols and equipment, this subtle difference may be explained by the distinct constructs measured between studies. While related, peak force (used in our study) is the maximal linear force produced during a task, whereas peak torque (used by McNabb et al (30)) is the maximal rotational force produced during a task (43). Population differences may have also contributed, as we studied recreational cyclists, rather than a general cohort of healthy adults.

As this is the first study to evaluate the Forceframe Max for hip flexion, direct comparisons to other studies are not possible. A previous meta-analysis reported that externally stabilised dynamometers in supine (as tested in our study) had excellent intra-tester reliability for (WCC = 0.93, 95%CI = 0.89-0.98; average SEMs = 35.50N) (27). We found that within session FF reliability was consistent with these findings (ICC_3.1_ = 0.89-0.95), but between-session reliability was slightly lower (ICCs_3.k=_ 0.77-0.84). However, the FF offered greater precision both within and between-sessions (SEM = 7.80 – 18.98N (3.47-8.54%)). This may reflect differences in the devices included in the meta-analysis, which consisted of various B-HHDs and other dynamometers that may not be directly comparable with the FF.

Our results closely aligned with those reported for hip adduction (test-retest ICC_2.1_= 0.81-0.90; SEM= 6.4-8.6%)) and adduction (test-retest ICC _2.1_= 0.72-0.92 SEM =4.3-11.9%) peak force measurment, using a different iteration of the FF in field sport athletes (33). These findings tentatively suggest that different FF models may perform consistently across populations and hip muscle groups, though further studies are needed to confirm this.

For hip extension, both devices demonstrated good to excellent reliability and similar measurement error (B-HHD ICCs = 0.90-0.95, SEMs = 15.77-21.53N (or 7.38-9.96%); FF ICCs = 0.85-0.95, SEMs =19.21-29.05N (7.82-11.78%) within and between sessions). These were comparable with values previously reported in a previous meta-analysis of externally stabilised dynamometers used in prone positions ((WCC = 0.90; 95%CI = 0.66-0.99; average SEM = 16.90N) (27). Similarly to hip flexion, we also found B-HHD reliability was consistent with values reported by McNabb et al. (30) for peak torque (ICC_2.1_ = 0.95; SEM= 12.00%), although SEM were marginally less in our study. (30).

As before, the FF cannot be compared with previous hip extension data, though our observed values were similar to those previously discussed, from a different Forceframe iteration for hip abduction and abduction peak force. (33).

Collectively, these findings suggest that while both devices in our study are reliable for hip extension MVIC peak force measurement, the close consistency with other dynamometers, muscle groups and measurement parameters may be a function of the stable prone test position for hip extension, but this should be also be confirmed in future studies.

### Minimal detectable change and clinical implications

Our observed MDC values for both devices suggest potential practical limitations for both devices. For hip flexion, the FF demonstrated consistently lower within-session MDCs (FF= 21.61N-39.48N (9.6-18.00%); B-HHD=39.50-62.98N (20.0-28.8%)). However, between-session MDCs were similar (MDC range=41.21-52.61N) and generally consistent with those reported for all externally stabilised portable dynamometers (MDC = 46.7N-49.3N) (27), but slightly less than 32% observed for peak torque measurement by McNabb et al (30) using a B-HHD For hip extension,the MDCs observed for both devices (MDCs= 43.71-67.12N ( 20.50-27.80%) were also broadly comparable with these previous reports (27, 30).

MDC values are crucial for distinguishing true performance changes from measurement error (40). Considering that strength changes of less than 15% are clinically and practically important (26), all within-session B-HHD MDCs, and three quarters of FF MDCs were greater than 15% of mean values. Furthermore, for both devices, all between-session MDCs were greater than 15% of mean values (range= 18.50% to 32.60%). This raises questions about the sensitivity of both devices to detect small but important true changes in performance within-session (e.g. to quantify the immediate effect of physiotherapy treatment, such as manual therapy or proprioceptive taping interventions), or over time as a response to rehabilitation or resistance training.

For illustration, isometric training may result in strength improvements of 4.34% per week (44), and 21.5% after 6-12 weeks (45) in healthy adults. Dynamic resistance training can result in 1.77% (46) to 5.20% (47) improvements in strength per week, increasing by 27.00% to 32.00% after 6 weeks (45, 46, 48).

Because these weekly adaptations are considerably less than all observed MDC values, it is unlikely that either device could detect true peak force changes if measured on a week to week basis. Furthermore, the similarity between six-week adaptations and between-session MDC estimates suggests that that any longer term (i.e >6 week) changes should also be cautiously interpreted as these may still be influenced by measurement error.

### Between system agreement

Despite the established concurrent validity of the MicroFET2 with the Kanagtech KT360, (42), a similar device to the FF, the poor agreement between devices for hip flexion and extension was surprising, especially considering the magnitude by which the 95%LOA were exceeded. Possible reasons may include differences in device resolution (FF =1.00N; MicroFET B-HHD= 0.44N) (49, 50), testing positions (i.e. the seated B-HHD hip flexion position my be less stable than the supine FF position), and the greater stability provided by the FF’s rigid aluminium frame and base, versus the B-HHD’s fabric belt.

Systematic bias for hip flexion was low (mean agreement range = −0.88-8.22 N). However, there was consistent bias for hip extension, with the FF recording higher peak force (28.04 to 45.38 N). Importantly, across all measurement scenarios presented in Figs 1 and 2, even adjusting values by the largest recorded mean difference (45.38N), the LOA would still exceed the 20N acceptability threshold, making this an unhelpful transformation.

This means that hip flexion or extension peak force data cannot be interchanged between devices. Measurements should therefore be considered as device-specific, so the same device should be used for repeated measurements.

### Limitations

Our study has some limitations. The marginal increase in peak force between days suggests a minor learning effect despite familiarisation. While no evidence of fatigue was observed, the validated Rating of Fatigue Scale (51), or other subjective wellbeing scores (52) could have quantified fatigue effects. The second test session was conducted approximately 72 hours after the first, but there was some variability around this due to participant’s external commitments, so could be most robustly controlled for in future studies.

Finally, since we used two-way mixed effects (ICC 3.1 and 3.k) models, our findings are not generalisable (38, 53). Future studies should replicate and confirm our data across different populations. This includes elite cylists, who are likely to utilise regular strength assessments.

## Conclusion

This study found that that the MicroFET2 and ForceFrame Max have good to excellent reliability and comparable precision for assessing hip flexor and extensor peak force in recreational cyclists. However, their MDC values limit clinical or practical usefulness in this population. The poor agreement prevents interchangeability between devices. Given the two-way mixed effects ICC models used, further studies are recommended in various populations to confirm these findings.

## Data Availability

All relevant data are within the manuscript and its Supporting Information files.

## Acknowledgements

The authors would like to thank all staff within the Manchester Metropolitan University Sport team, and British Cycling Clubs and Community Groups for their help and support with participant recruitment (without whom this study would not be possible). The authors also thank: Katie Flatters and Heather Stephens at British Cycling for support with study conception; Prof. Michael Callaghan at Manchester Metropolitan University for methodology advice; Dr. Jamie Sergeant at University of Manchester for statistical advice, and; Liam Parkinson from VALD Performance for providing the VALD ForceFrame Max.

## Author Contributions

Conceptualisation: DDM, BD, TH.

Data curation: DDM, TH.

Formal analysis: DDM, TH.

Investigation: DDM.

Methodology: DDM, BD, TH.

Project administration: DDM.

Resources: DDM.

Software: DDM, TH.

Supervision: TH.

Visualisation: DDM, BD, TH.

Writing – Original draft and preparation: DDM.

Writing – review and editing: DDM, BD, TH.

## Supporting Information Captions

S1 Appendix. Sample size calculation

S2 Appendix. Testing positions.

S3 Appendix. Quantile quantile plots.

S4 Appendix. Limits of agreement estimates.

## References

1. Oja P, Titze S, Bauman A, De Geus B, Krenn P, Reger-Nash B, et al. Health benefits of cycling: a systematic review. Scandinavian Journal of Medicine & Science in Sports. 2011;21(4):496–509.

2. Leyland L-A, Spencer B, Beale N, Jones T, Van Reekum CM. The effect of cycling on cognitive function and well-being in older adults. PloS One. 2019;14(2):e0211779.

3. Nordengen S, Andersen LB, Solbraa AK, Riiser A. Cycling is associated with a lower incidence of cardiovascular diseases and death: Part 1–systematic review of cohort studies with meta-analysis. British Journal of Sports Medicine. 2019;53(14):870–8.

4. National Institute of Health and Care Excellence (NICE). Walking and cycling: local measures to promote walking and cycling as forms of travel or recreation [PH41] 2012 [updated 28 February 2019. Available from: https://www.nice.org.uk/guidance/ph41.

5. Pucher J, Buehler R, Seinen M. Bicycling renaissance in North America? An update and re-appraisal of cycling trends and policies. Transportation Research Part A: Policy and Practice. 2011;45(6):451–75.

6. Jancaitis G, Snyder Valier AR, Bay C. A descriptive and comparative analysis of injuries reported in USA Cycling-sanctioned competitive road cycling events. Injury Epidemiology. 2022;9(1):22.

7. Buehler R, Pucher J, Merom D, Bauman A. Active travel in Germany and the US: contributions of daily walking and cycling to physical activity. American Journal of Preventive Medicine. 2011;41(3):241–50.

8. Department for Transport. Road Traffic Estimates: Great Britain 2021. Annual Road Traffic Estimates Statistical Release. London: Department for Transport; 2021.

9. Dorel S, Couturier A, Lacour J-R, Vandewalle H, Hautier C, Hug F. Force-velocity relationship in cycling revisited: benefit of two-dimensional pedal forces analysis. Medicine and Science in Sports and Exercise. 2010;42(6):1174–83.

10. Martin-Rodriguez S, Gonzalez-Henriquez JJ, Bautista IJ, Calbet JA, Sanchis-Moysi J. Interplay of muscle architecture, morphology, and quality in influencing human sprint cycling performance: A systematic review. Sports Medicine-Open. 2024;10(1):81.

11. So RC, Ng JK-F, Ng GY. Muscle recruitment pattern in cycling: a review. Physical Therapy in Sport. 2005;6(2):89–96.

12. Yamamoto LM, Klau JF, Casa DJ, Kraemer WJ, Armstrong LE, Maresh CM. The effects of resistance training on road cycling performance among highly trained cyclists: a systematic review. The Journal of Strength & Conditioning Research. 2010;24(2):560–6.

13. Bastiaans J, Diemen A, Veneberg T, Jeukendrup A. The effects of replacing a portion of endurance training by explosive strength training on performance in trained cyclists. European Journal of Applied Physiology. 2001;86(1):79–84.

14. Rønnestad BR, Hansen EA, Raastad T. Effect of heavy strength training on thigh muscle cross-sectional area, performance determinants, and performance in well-trained cyclists. European Journal of Applied Physiology. 2010;108:965–75.

15. Vikmoen O, Rønnestad BR. A comparison of the effect of strength training on cycling performance between men and women. Journal of Functional Morphology and Kinesiology. 2021;6(1):29.

16. Rønnestad BR, Hansen J, Hollan I, Ellefsen S. Strength training improves performance and pedaling characteristics in elite cyclists. Scandinavian Journal of Medicine & Science in Sports. 2015;25(1):e89–e98.

17. Hansen EA, Raastad T, Hallén J. Strength training reduces freely chosen pedal rate during submaximal cycling. European Journal of Applied Physiology. 2007;101:419–26.

18. Clarsen B, Pluim BM, Moreno-Pérez V, Bigard X, Blauwet C, Del Coso J, et al. Methods for epidemiological studies in competitive cycling: an extension of the IOC consensus statement on methods for recording and reporting of epidemiological data on injury and illness in sport 2020. British Journal of Sports Medicine. 2021;55(22):1262–9.

19. Hughes T, Sergeant JC, van der Windt DA, Riley R, Callaghan MJ. Periodic health examination and injury prediction in professional football (soccer): theoretically, the prognosis is good. Sports Medicine. 2018;48(11):2443–8.

20. Jamnick NA, Pettitt RW, Granata C, Pyne DB, Bishop DJ. An examination and critique of current methods to determine exercise intensity. Sports Medicine. 2020;50(10):1729–56.

21. Suchomel TJ, Nimphius S, Stone MH. The importance of muscular strength in athletic performance. Sports Medicine. 2016;46:1419–49.

22. Stark T, Walker B, Phillips JK, Fejer R, Beck R. Hand-held dynamometry correlation with the gold standard isokinetic dynamometry: a systematic review. PM&R. 2011;3(5):472–9.

23. Desmyttere G, Gaudet S, Begon M. Test-retest reliability of a hip strength assessment system in varsity soccer players. Physical Therapy in Sport. 2019;37:138–43.

24. Martins J, Da Silva JR, Da Silva MRB, Bevilaqua-Grossi D. Reliability and validity of the belt-stabilized handheld dynamometer in hip-and knee-strength tests. Journal of Athletic Training. 2017;52(9):809–19.

25. Jackson SM, Cheng MS, Smith Jr AR, Kolber MJ. Intrarater reliability of hand held dynamometry in measuring lower extremity isometric strength using a portable stabilization device. Musculoskeletal Science and Practice. 2017;27:137–41.

26. Chamorro C, Armijo-Olivo S, De la Fuente C, Fuentes J, Javier Chirosa L. Absolute reliability and concurrent validity of hand held dynamometry and isokinetic dynamometry in the hip, knee and ankle joint: systematic review and meta-analysis. Open Medicine. 2017;12(1):359–75.

27. Waiteman MC, Garcia MC, Briani RV, Norte G, Glaviano NR, De Azevedo FM, et al. Can Clinicians Trust Objective Measures of Hip Muscle Strength From Portable Dynamometers? A Systematic Review With Meta-analysis and Evidence Gap Map of 107 Studies of Reliability and Criterion Validity Using the COSMIN Methodology. Journal of Orthopaedic and Sports Physical Therapy. 2023;53(11):655–72.

28. Thorborg K, Bandholm T, Schick M, Jensen J, Hölmich P. Hip strength assessment using handheld dynamometry is subject to intertester bias when testers are of different sex and strength. Scandinavian Journal of Medicine & Science in Sports. 2013;23(4):487–93.

29. Kolber MJ, Cleland JA. Strength testing using hand-held dynamometry. Physical Therapy Reviews. 2005;10(2):99–112.

30. McNabb K, Sánchez MB, Selfe J, Reeves ND, Callaghan M. Handheld dynamometry: Validity and reliability of measuring hip joint rate of torque development and peak torque. Plos One. 2024;19(8):e0308956.

31. Mokkink LB, Terwee CB, Patrick DL, Alonso J, Stratford PW, Knol DL, et al. The COSMIN checklist for assessing the methodological quality of studies on measurement properties of health status measurement instruments: an international Delphi study. Quality of Life Research. 2010;19:539–49.

32. Kottner J, Audigé L, Brorson S, Donner A, Gajewski BJ, Hróbjartsson A, et al. Guidelines for reporting reliability and agreement studies (GRRAS) were proposed. International Journal of Nursing Studies. 2011;48(6):661–71.

33. O’Connor C, McIntyre M, Delahunt E, Thorborg K. Reliability and validity of common hip adduction strength measures: the ForceFrame strength testing system versus the sphygmomanometer. Physical Therapy in Sport. 2023;59:162–7.

34. Ransom M, Saunders S, Gallo T, Segal J, Jones D, Jones M, et al. Reliability of a portable fixed frame dynamometry system used to test lower limb strength in elite Australian Football League players. Journal of Science and Medicine in Sport. 2020;23(9):826–30.

35. Gagnier JJ, Lai J, Mokkink LB, Terwee CB. COSMIN reporting guideline for studies on measurement properties of patient-reported outcome measures. Quality of Life Research. 2021;30:2197–218.

36. D’Mello D, Digweed B, Hughes T. Test-retest reliability and agreement of a Belt-stabilized Handheld Dynamometer and the ForceFrame Strength testing system in hip strength testing of recreational cyclists. 2024. Open Science Framework. 10.17605/OSF.IO/8HB7M.

37. Brown W, Naughton G, Oldenburg B, Owen N, Wright MC. Promoting physical activity—: Atlanta: Centers for Disease Control and Prevention; 2010.

38. Mokkink LB, de Vet H, Diemeer S, Eekhout I. Sample size recommendations for studies on reliability and measurement error: an online application based on simulation studies. Health Services and Outcomes Research Methodology. 2023;23(3):241–65.

39. Coppieters M, Stappaerts K, Janssens K, Jull G. Reliability of detecting ‘onset of pain’and ‘submaximal pain’during neural provocation testing of the upper quadrant. Physiotherapy Research International. 2002;7(3):146–56.

40. Weir JP. Quantifying test-retest reliability using the intraclass correlation coefficient and the SEM. The Journal of Strength & Conditioning Research. 2005;19(1):231–40.

41. Bland JM, Altman D. Statistical methods for assessing agreement between two methods of clinical measurement. The Lancet. 1986;327(8476):307–10.

42. Dunne C, Callaway AJ, Thurston J, Williams JM. Validity, reliability, minimal detectable change, and methodological considerations for HHD and portable fixed frame isometric hip and groin strength testing: A comparison of unilateral and bilateral testing methods. Physical Therapy in Sport. 2022;57:46–52.

43. Garcia MAC, Fonseca DS, Souza VH. Handheld dynamometers for muscle strength assessment: Pitfalls, misconceptions, and facts. Brazilian Journal of Physical Therapy. 2020;25(3):231.

44. Oranchuk DJ, Storey AG, Nelson AR, Cronin JB. Isometric training and long-term adaptations: Effects of muscle length, intensity, and intent: A systematic review. Scandinavian Journal of Medicine & Science in Sports. 2019;29(4):484–503.

45. Schoenfeld BJ, Grgic J, Ogborn D, Krieger JW. Strength and hypertrophy adaptations between low-vs. high-load resistance training: a systematic review and meta-analysis. The Journal of Strength & Conditioning Research. 2017;31(12):3508–23.

46. Moesgaard L, Beck MM, Christiansen L, Aagaard P, Lundbye-Jensen J. Effects of periodization on strength and muscle hypertrophy in volume-equated resistance training programs: a systematic review and meta-analysis. Sports Medicine. 2022;52(7):1647–66.

47. Jung R, Gehlert S, Geisler S, Isenmann E, Eyre J, Zinner C. Muscle strength gains per week are higher in the lower-body than the upper-body in resistance training experienced healthy young women—A systematic review with meta-analysis. PloS one. 2023;18(4):e0284216.

48. Hagstrom AD, Marshall PW, Halaki M, Hackett DA. The effect of resistance training in women on dynamic strength and muscular hypertrophy: a systematic review with meta-analysis. Sports Medicine. 2020;50(6):1075–93.

49. Scientific H. microFET2 Digital Handheld Dynamometer [Internet]. Salt Lake City (UT): Hoggan Scientific; 2023 [Available from: https://hogganscientific.com/product/microfet2-muscle-tester-digital-handheld-dynamometer/

50. VALD. ForceFrame Technical Specifications [Internet]. Brisbane (AU): VALD; 2023 [updated 17 Nov 2023. Available from: https://support.vald.com/hc/en-au/articles/4998527361049-ForceFrame-Technical-Specifications.

51. Micklewright D, St Clair Gibson A, Gladwell V, Al Salman A. Development and validity of the rating-of-fatigue scale. Sports Medicine. 2017;47:2375–93.

52. Saw AE, Main LC, Gastin PB. Monitoring the athlete training response: subjective self-reported measures trump commonly used objective measures: a systematic review. British Journal of Sports Medicine. 2016;50(5):281–91.

53. Koo TK, Li MY. A guideline of selecting and reporting intraclass correlation coefficients for reliability research. Journal of Chiropractic Medicine. 2016;15(2):155–63.

